# Three-month outcomes in hospitalized COVID-19 patients

**DOI:** 10.1101/2020.10.16.20211029

**Authors:** Jude PJ Savarraj, Angela B Burkett, Sarah N Hinds, Atzhiry S Paz, Andres Assing, Shivanki Juneja, Gabriela Delevati Colpo, Luis F Torres, Aaron Gusdon, Louise McCullough, H Alex Choi

## Abstract

COVID-19 is an ongoing pandemic with a devastating impact on public health. Acute neurological symptoms have been reported after a COVID-19 diagnosis, however there is no data available on the long-term neurological symptoms. Using a prospective registry of hospitalized COVID-19 patients, we assessed the neurological assessments (including functional, cognitive and psychiatric assessments) of several hospitalized patients at 3 months. Our main finding is that 71% of the patients still experienced neurological symptoms at 3 months and the most common symptoms being fatigue (42%) and PTSD (29%). 64% of the patients report pain symptoms we well. Cognitive symptoms were found in 12%. Our preliminary findings suggests the importance of investigating long-term and rationalizes the need for further studies investigating the neurologic outcomes after COVID-19.

## Introduction

To date over 36 million people have been infected with the *severe acute respiratory syndrome coronavirus 2* virus (SARS-CoV-2), which causes the coronavirus disease 2019 (COVID-19). While the vast majority will survive, many may be left with residual effects. Acute neurologic symptoms after COVID-19 including encephalitis, acute myopathic quadriplegia, strokes and seizures have been reported.^1^ Anecdotal reports of long-term neurologic symptoms are emerging as well.^2^ These reports have emphasized the importance of studying long-term neurologic outcomes, also referred as the “Long-Haul COVID”. ^3^ Short- and long-term neurologic symptoms were reported during the SARS and MERS outbreaks^4^ and it is likely that long-term symptoms will persist after COVID-19 as well. To characterize long-term neurologic outcomes after COVID-19 we followed a cohort of hospitalized patients and assessed 3-months outcomes.

## Methods

We conducted a prospective single-center study of hospitalized COVID-19 patients admitted to the University of Texas Health Science Center at Houston, Texas from May 2020 to July 2020 during the surge seen in Texas, USA. Inclusion criteria were laboratory-confirmed SARS-CoV-2 infection by real-time PCR and admission to the hospital for COVID-19. All hospitalized patients were either hospitalized mild (WHO scale 4, requiring oxygen by mask or nasal cannula) or severe (WHO scale ≥ 5, requiring at least high–flow oxygen)^5^. Exclusion criteria were subjects with diagnosis of pre-morbid conditions interfering with outcome domains being assessed. Written informed consent was obtained from subject or legal surrogate. This study was approved by the Institutional Review Board, No: HSC-MS-20-1011. 140 subjects were enrolled. 65 were lost to follow-up and 27 were dead. 3-month outcomes were determined in 48 subjects using telephone questionnaires to assess functional, cognitive and psychiatric symptoms. Functional outcome was evaluated using the modified Rankin Score (mRS). Cognitive status was evaluated using the brief neurocognitive screening test (BNST). Depression symptoms were evaluated using the Patient Health Questionnaire (PHQ-9). Anxiety symptoms were assessed using the Generalized Anxiety Disorder (GAD-7). Pain, fatigue and sleepiness were evaluated using the Pain, Enjoyment of life and General activity (PEG), the Fatigue Severity Scale (FSS) and Epworth Sleepiness Scale (ESS). Post-traumatic stress disorder was evaluated using the Primary Care PTSD Screen for DSM-5 (PC-PTSD-5).

## Results/Discussion

In our cohort of hospitalized COVID-19 survivors, 71% had continued neurologic symptoms highlighting the importance of considering the *long-haul COVID* phenomena. The most common symptom was fatigue (42%) followed by PTSD symptoms (29%). [Table 1] People with long-term symptoms were significantly older (mean, years (SD): 54 (16) v 41 (16); *p=0*.*01*). The persistence of long-term symptoms was not associated with the severity of acute COVID-19 symptoms. Neither the maximum C-reactive protein levels [(137 (73) v 153 (92); *p=0*.*59*] nor the clinical severity [WHO≤4 v WHO>5, *p=0*.*58*] were associated with 3-month symptoms. In fact, we found that even subjects with mild course of hospitalization had a high incidence of symptoms, especially fatigue (58%). Our findings support the anecdotal reports of long-term symptoms even in those with mild acute pulmonary symptoms. The prevalence of cognitive symptoms (12%) were relatively lower than generalized symptoms. However, 12% of survivors represents a large number of people altogether, and gives credence to the reports of “brain fog” that survivors have experienced. Although symptoms of pain were frequent (64%), there is no standardized cut off for pain measurements so was not included in our analysis of combined neurologic symptoms. In those subjects who described pain, the PEG score was 4.26 ± 2.7 (mean ± SD), a score similar to ones reported in other diseases in which pain is prevalent after hospital discharge^6^.

**Table:**

| **Demographics** | **N = 48** |  |
| --- | --- | --- |
| Age (mean, sd) | 50(17) |  |
| Sex (Female, %) | 23(48) |  |
| Ethnicity (Hispanic, %) | 32(67) |  |
| **Past Medical History** |  |  |
| Obesity (n, %) | 25(52) |  |
| Diabetes | 18(38) |  |
| Hypertension | 24(50) |  |
| Smoking (n, %) | 5(11) |  |
| COPD | 2(4) |  |
| CCI (median, IQR) | 2[1-3] |  |
| WHO Classification (Severe, %) | 21(44) |  |
| **Outcomes** | **N,%** | **Responses** |
| Any neurologic symptom | 34(71%) | 48 |
| Fatigue symptoms (FSS) | 19(42%) | 45 |
| Post-traumatic stress disorder (PC-PTSD-5) | 12(29%) | 44 |
| Functional outcome (mRS) | 10(21%) | 48 |
| Sleepiness (ESS) | 7(17%) | 42 |
| Cognitive Deficit (BNST) | 5(12%) | 43 |
| Depression Symptoms (PHQ-9) | 5(11%) | 45 |
| Anxiety (GAD-7) | 4(9%) | 45 |
| Pain (PEG) | 29(64%) | 45 |

The mRS is a 0-6 scale with a cutoff of $\geq 3$ indicative of functional disability. BNST is a 0-12 scale with a cutoff of $\leq 8$ indicative of cognitive symptoms. The PHQ-9 is a 0-20 with a cutoff of $\geq 10$ indicative of depression symptoms. The GAD-7 is a 0-21 with a cutoff of $\geq 10$ is indicative of general anxiety syndrome. PEG is a 0-10 point scale with no cutoff. FSS is a a 0-7 point scale with a cutoff of $\geq 4$ indicative of fatigue and ESS is a 0-24 point scale with a cutoff of $\geq 11$ indicative of sleepiness symptoms. The PC-PTSD-5 is a 0-5 point scale with a cutoff of $\geq 3$ indicative of PTSD).

Despite the small sample size, this is one of the first study to prospectively evaluate long-term neurologic effects seen COVID-19 patients. All the assessments were performed using phone questionnaires instead of in-person assessments. However, remote contactless assessments are necessary as the pandemic has changed the care paradigm (causing disruption in routine health care services) and many recovering patients are reluctant to participate in an in-person follow-up due to health limitations. Since the population was limited to a hospitalized patient cohort the results cannot be extrapolated to milder formed of COVID-19 that did not require hospitalization. The influence of age and co-morbidities were not analyzed due to the small sample size. Furthermore, this study does not include a suitable control cohort.

Although studies have reported acute neurological symptoms after COVID-19, our study is one of the first to examine the persistence of neurologic symptoms at 3-months. Studies examining pathophysiology and the time course of persistent neurologic symptoms after COVID-19 are needed. Our findings emphasize the importance of continued evaluation and focused rehabilitation for functional, cognitive and neurobehavioral consequences in COVID-19 survivors.

## Data Availability

Data available upon request.

## Acknowledgements

The authors thank all the health care workers and the research personnel who were involved in the care and the follow-up of the patients.

## Appendix Authors

| Name | Location | Role | Contribution |
| --- | --- | --- | --- |
| Jude PJ Savarraj, PhD | Department of Neurosurgery,<br>McGovern Medical School<br>The University of Texas Health<br>Science Center at Houston<br>Texas, USA | Author | Study concept and design<br>Critical revision of manuscript<br>for intellectual content |
| Angela Burkett, BS | Department of Neurosurgery,<br>McGovern Medical School<br>The University of Texas Health<br>Science Center at Houston<br>Texas, USA | Author | Study concept, data collection<br>Critical revision of manuscript<br>for intellectual content |
| Sarah Hinds, BS | Department of Neurosurgery,<br>McGovern Medical School<br>The University of Texas Health<br>Science Center at Houston<br>Texas, USA | Author | Study concept, data collection<br>Critical revision of manuscript<br>for intellectual content |
| Atzhiry Paz, BS | Department of Neurosurgery,<br>McGovern Medical School<br>The University of Texas Health<br>Science Center at Houston<br>Texas, USA | Author | Study concept, data collection<br>Critical revision of manuscript<br>for intellectual content |
| Andres Assing, BS | Department of Neurosurgery,<br>McGovern Medical School<br>The University of Texas Health<br>Science Center at Houston<br>Texas, USA | Author | Study concept, data collection<br>Critical revision of manuscript<br>for intellectual content |
| Shivanki Juneja, BS | Department of Neurology,<br>McGovern Medical School<br>The University of Texas Health<br>Science Center at Houston<br>Texas, USA | Author | Study concept, data collection<br>Critical revision of manuscript<br>for intellectual content |
| Gabriela Delevati Colpo, PhD | Department of Neurology,<br>McGovern Medical School<br>The University of Texas Health<br>Science Center at Houston<br>Texas, USA | Author | Study concept, data collection<br>Critical revision of manuscript<br>for intellectual content |
| Luis Torres, MD | Department of Neurosurgery,<br>McGovern Medical School<br>The University of Texas Health<br>Science Center at Houston<br>Texas, USA | Author | Study concept and design<br>Critical revision of manuscript<br>for intellectual content |
| Aaron Gusdon, MD | Department of Neurosurgery,<br>McGovern Medical School<br>The University of Texas Health<br>Science Center at Houston<br>Texas, USA | Author | Study concept and design<br>Critical revision of manuscript<br>for intellectual content |
| Louise McCullough, MD, PhD | Department of Neurology,<br>McGovern Medical School<br>The University of Texas Health<br>Science Center at Houston<br>Texas, USA | Author | Study concept and design<br>Critical revision of manuscript<br>for intellectual content |
| H Alex Choi, MD | Department of Neurosurgery,<br>McGovern Medical School<br>The University of Texas Health<br>Science Center at Houston<br>Texas, USA | Author | Study concept and design<br>Critical revision of manuscript<br>for intellectual content |

